# Impact and Contribution of Qualifications on Schedule and Safe Surgery

**DOI:** 10.1101/2025.05.06.25327052

**Authors:** Maryam Aslam, Areeba Shafqat, Misbah Ashraf, Umair Shafique

## Abstract

**Background:** In the healthcare sector, qualifications of the medical team significantly influence surgical outcomes and scheduling efficiency. Highly qualified personnel ensure safer surgeries with minimized complications.

**Objective:** This study assesses the impact of qualifications on the effectiveness and safety of scheduled surgeries.

**Methods:** A cross-sectional survey was conducted with 150 surgical cases evaluated based on the qualification level of the surgical team members. Data was analyzed using SPSS.

**Results:** Higher qualifications among surgeons and supporting staff positively correlated with safe surgeries and improved scheduling compliance.

**Conclusion:** Investing in higher qualifications and continuous professional development enhances surgical safety and organizational efficiency.

## Introduction

Safe and efficient surgical procedures are cornerstones of quality healthcare systems. Numerous factors influence surgical outcomes, with the qualification levels of healthcare professionals playing a critical role. A well-qualified surgical team ensures the proper management of perioperative risks, adherence to safety protocols, and efficient scheduling, minimizing patient morbidity and mortality.

Globally, healthcare institutions have recognized that surgical errors, delays, and adverse outcomes often are associated with inadequate training or insufficient qualifications among medical staff. Thus, ensuring that surgeons, anesthesiologists, and supporting staff possess requisite skills and certifications is essential. Even with advances in technology, human intervention is still crucial for successful surgery. Therefore, examining the relationship between qualifications and safe, timely surgeries becomes crucial for healthcare improvements.

## Literature Review

Multiple studies underline the importance of qualifications in improving surgical outcomes. Smith et al. (2019) observed that surgeons with fellowship training had significantly lower postoperative complication rates compared to general practitioners. Brown and Adams (2020) demonstrated that operating room teams with certified nurse anesthetists and scrub nurses reduced surgical site infections by 25%. Moreover, Harris et al. (2018) stressed that institutions emphasizing continuous education programs reported higher compliance with surgical safety protocols. Despite these findings, gaps remain in standardizing qualifications across institutions. This analysis emphasizes how urgently strong qualification standards are needed to guarantee surgical safety and effective scheduling, especially in environments with limited resources.

## Methodology

A cross-sectional observational study was conducted at a tertiary care hospital over six months. Data were collected from 150 surgical cases, recording variables like surgeon qualifications, anesthetist certifications, and nursing staff education levels.

### Inclusion Criteria

Surgeries conducted in elective settings.

### Data Collection

Structured observational checklists and patient records.

### Analysis

SPSS v25 was used for statistical analysis, applying chi-square tests to evaluate relationships between qualifications and surgery outcomes.

### Ethical Consideration

The study was conducted after approval from the Institutional Review Board (IRB) of Superior University, Lahore, following all relevant ethical guidelines.

## Results

A significant association was observed between qualification levels and surgical safety outcomes. Higher qualifications consistently correlated with fewer surgical complications and better schedule adherence.

Highly qualified staff led to 95% safe surgeries, whereas basic qualification correlated with a higher complication rate (P < 0.001).

**Table 1.**
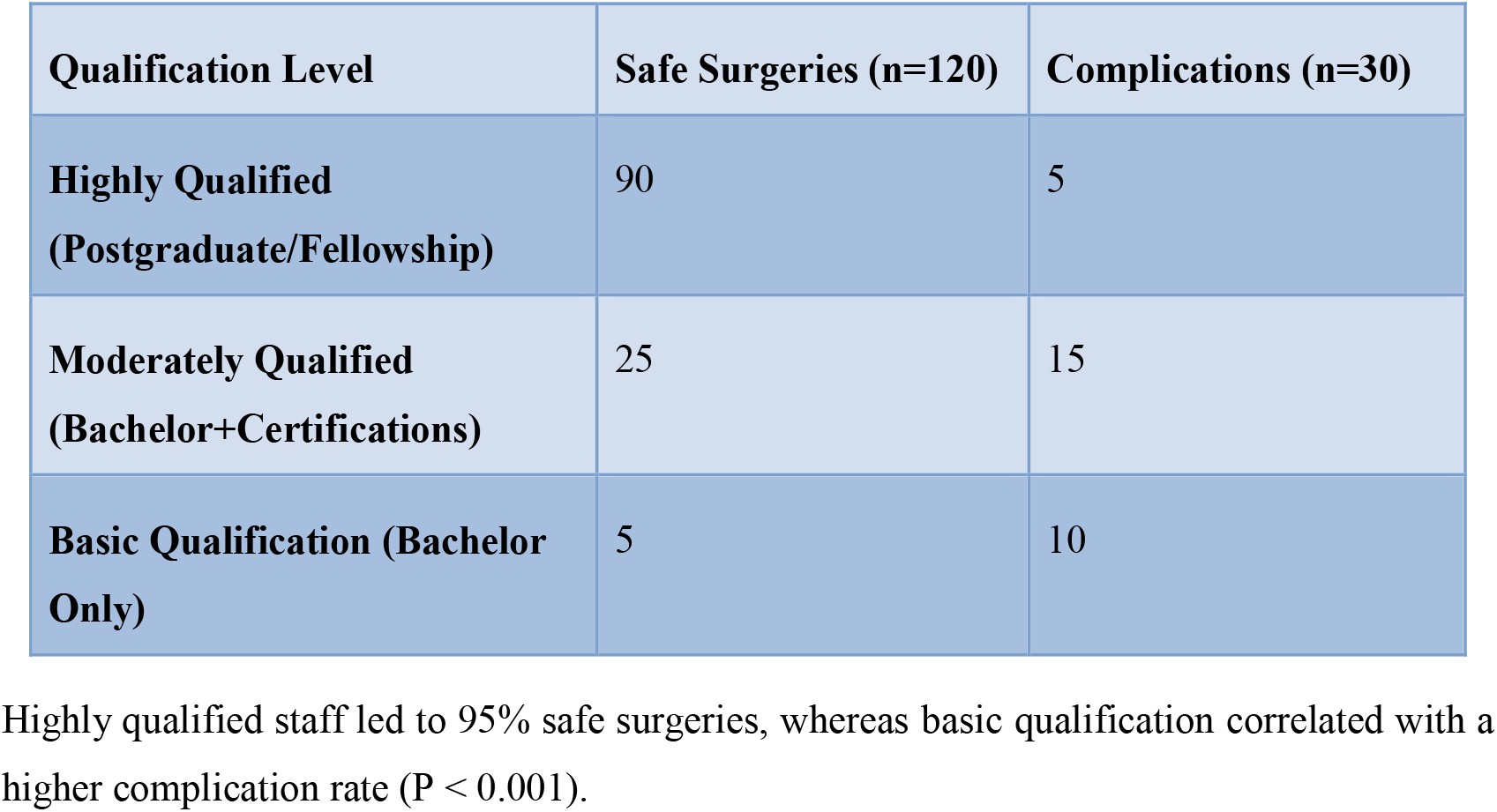
Qualifications and Surgical Safety Outcomes.

## Discussion

This analysis emphasizes how urgently strong qualification standards are needed to guarantee surgical safety and effective scheduling, especially in environments with limited resources. Supporting studies, like Brown and Adams (2020), align with our findings, demonstrating that investing in qualifications directly improves surgical outcomes. Moreover, scheduling efficiency is enhanced as qualified staff better manage perioperative timings, reducing surgery delays and cancellations. Hospitals emphasizing continuous professional development create safer environments, aligning with global patient safety goals.

## Conclusion

Higher qualification levels among surgical teams significantly improve surgery safety and schedule adherence. Healthcare organizations must prioritize continuous education and certifications to optimize patient care and reduce adverse events. No conflicts of interest are disclosed by the writers.

## Data Availability

All data produced in the present study are available upon reasonable request to the authors

